# Applicability of a short form of the Speech, Spatial and Qualities of Hearing Scale in 97 individuals with Menière’s disease in a multicenter registry

**DOI:** 10.1101/2025.01.01.25319849

**Authors:** Jennifer L. Spiegel, Bernhard Lehnert, Laura Schuller, Irina Adler, Tobias Rader, Tina Brzoska, Bernhard G. Weiss, Martin Canis, Chia-Jung Busch, Friedrich Ihler

## Abstract

**BACKGROUND:** Menière’s Disease (MD) is a debilitating disorder with episodic and variable ear symptoms. Diagnosis can be challenging and there is no current consensus regarding monitoring of disease progression. A psychometric instrument may aid the assessment of subjective hearing impairment and the associated burden in daily life over the course of the disease. However, evidence in this regard is lacking.

**OBJECTIVE:** To evaluate a German-language short form of the Speech, Spatial and Qualities of Hearing Scale (SSQ) for hearing-related disability in MD.

**METHODS:** Data was collected from a multicenter prospective patient registry for long-term follow up of MD patients. Hearing was assessed by pure tone and speech audiometry. The applied version of the SSQ contained 17 items.

**RESULTS:** 97 consecutive patients with unilateral MD had a mean age of 56.2 ± 5.0 years. 55 individuals (57.3%) were female, 72 (75.0%) were categorized as definite MD. Average total score of the SSQ was 6.0 ± 2.1. Cronbach’s alpha for internal consistency was 0.960 for the total score. We did not observe undue floor or ceiling effects. SSQ values showed a negative correlation with hearing thresholds and a positive correlation with speech recognition scores.

**CONCLUSIONS:** The short form of the SSQ provides insight into hearing-specific disability in patients with MD. Thereby, it may be informative regarding disease stage and rehabilitation needs.

## Introduction

Menière’s Disease (MD) is characterized by recurrent episodes of vertigo or dizziness, hearing loss and other aural symptoms. Diagnosing MD can be complicated by the episodic and fluctuating nature of the disease; therefore the *Barany Society* has introduced internationally consented diagnostic criteria. By considering the duration of vertigo attacks and audiometric documentation of fluctuating low-frequency hearing loss, patients are assigned to two categories of diagnostic certainty: definite and probable MD [31]. However, those criteria may not capture the whole clinical picture of the disease [25], hence making a diagnosis challenging in many cases. With a full mechanistic explanation pending, potential biomarkers like endolymphatic hydrops, which can be visualized by contrast-enhanced MRI, still have to prove their clinical value [33,44,46]. While a range of diverse therapeutic options exists, most are supported by a low level of evidence [8,15].

In particular the recurring bouts of vertigo significantly debilitate patients in their daily life activities, work and private relationships [55]. The psychological repercussions of vestibular disorders are widely acknowledged [6,22] and can even result in aggravation and prolonging of dizziness [39]. Intrinsically, psychological factors are linked to quality of life and *vice versa*, and in addition, there is even evidence for an associated economic burden [2].

Alongside recurrent vertigo episodes, patients experience hearing loss of fluctuating or permanent character. A prerequisite of categorizing patients according to current diagnostic criteria is assessing hearing loss. However, too little attention has been brought to evaluate hearing in MD patients so far [17,24] and in particular research focusing on hearing-related quality of life of MD patients is lacking. MD occurs unilaterally in the majority of cases [4,44] and consecutively, most MD patients develop asymmetric hearing loss, a condition that results in specific impairments in spatial tasks and challenging listening situations [30]. Data suggests that patients with unilateral profound hearing loss suffer a substantial burden of stress associated with increased hearing effort and a range of psychological and social consequences [34]. In social settings, patients feel the need to strategically position themselves in a group to optimize their ability to hear and participate [52]. Common associated struggles include restrictions within social life, and sometimes even exclusion, thus resulting in higher levels of handicap [14,26]. As per evaluating advanced hearing situations, the *Speech, Spatial and Quality of Hearing Scale* (SSQ) is a commonly applied and validated questionnaire, which was designed to capture the ability and impairment of hearing in daily life [18,37].

In order to gain a deeper understanding of hearing-loss-related quality of life in MD patients, available instruments are required to be evaluated upon feasibility in this specific cohort. Thus, we aim to explore whether the SSQ is a suitable tool to assess and measure hearing loss-related quality of life in unilateral MD.

## Material and Methods

### Study design, sites and ethical considerations

All patients are part of a multicenter prospective patient registry for MD (German title: “Systematische Erfassung von Morbus Menière: Prospektive Beobachtungsstudie”, acronym: “SEMM”). It was initiated in 2021 and is intended for long-term follow up. It is registered with the German Clinical Trial Registry under DRKS00027830. The study is approved by the research ethics boards (REB) of both study sites (REB approval number Faculty of Medicine of LMU Munich: 21-0779; REB approval number Greifswald University Medicine: BB 055/22), which are both tertiary academic centers with designated neurotological practice. The study was performed according to the current version of the Declaration of Helsinki.

### Patients and diagnosis

Patients were recruited at academic referral clinics for dizziness. All cases received comprehensive vestibular assessment, including clinical examination, videonystagmography, caloric testing, video head impulse test, and posturography. Retrocochlear causes were excluded by MRI with gadolinium of the brain with a focus on the cerebello-pontine-angle.

Patients were included if they fulfilled the criteria for MD according to the current diagnostic criteria, which comprise two diagnostic categories definite Menière’s disease (dMD) and probable Menière’s disease (pMD) [31]. To account for cases that presented with a clinical picture of typical Menière-symptoms but did not fit into those canonical categories, an additional diagnostic category was provided as described earlier (Menière’s Characteristics, MC) [25]. In all patients within this category, in case of central symptoms, e. g. leading to suspected vestibular migraine, a neurological consultation was performed.

Exclusion criteria for this analysis were vestibular schwannoma in imaging study, other vestibular disorder than Menière’s Disease, previous intracranial tumor or surgery, stroke, other central explanation for vestibular symptoms, and bilateral Menière’s Disease. Additional migraine symptoms to otherwise diagnosed MD were by itself no exclusion criterion.

### Questionnaire: The German-language SSQ short form (SSQ17)

The Speech, Spatial and Qualities of Hearing Scale (SSQ) [18] encompasses the three subscales speech hearing, spatial hearing and qualities of hearing, originally with a total of 49 questions. The questionnaire is available now in several versions with various numbers of questions and in different languages. Here, the only German version was applied in a short form with 17 items [27] to assess patient-reported hearing ability. This version comprises five items for each of the three subscales as well two additional questions on hearing in quiet and listening effort, respectively. A total score was calculated as mean and median of the results of all 17 items. Likewise, mean and median scores for the subscales were obtained. Across all items, higher numeric values signify higher hearing ability with a maximum of 10 while lower values represent increasing impairment with a minimum value of 0. Values were marked on a visual analogue scale and red out to one decimal. For each item individually, a checkbox was given to mark “not applicable” for the hearing situation in question.

### Pure-tone audiometry

Audiometric measurements were performed in quiet, soundproof, anechoic chambers by specialized and experienced technical staff. Pure tone thresholds were assessed for each ear separately from 0.5 to 8 kHz with air and bone conduction between -10 dB and 120 dB hearing level (dB HL) or up to uncomfortable loudness. A summary value of pure tone audiometry was calculated as average for the four air conducted frequencies 0.5, 1, 2, and 4 kHz (4PTA).

### Speech audiometry

Unaided speech recognition of monosyllabic words was assessed at 65 dB HL with the German language Freiburg Monosyllabic Test [21]. In brief, for each investigation a list of 20 monosyllabic words recorded from a male speaker was randomly chosen from a total of 20 lists. Monosyllabic words were consecutively presented to the patients for each ear separately via headphones. Each correctly recognized word was accounted for 5%, thereby the maximum score was 100%.

### Statistical analysis

Data was entered and pseudonymized in Microsoft Excel on site and merged and analyzed in R 4.4.1 (https://www.R-project.org/) [43]. Pure tone thresholds louder than measurable were set to 120 dB HL. The following imputations were performed: In one case a 4 kHz pure tone threshold was not available and replaced by this patients 3 kHz threshold for 4PTA computation, in three cases where monosyllable understanding was missing it was imputed via logistic regression from the 4PTA values, based on all other ears’ results.

The usual descriptive statistics are reported, using Gini mean difference (GMD) [20] over standard deviation for its advantages in measures, “in which the normal distribution and even a symmetric density function do not provide a good approximation to the data” [56]. Cronbach’s alpha [12] was used to assess internal consistency of the SSQ questionnaire’s scales. A 95%-Confidence Interval (95% CI) for Cronbach’s alpha was calculated as described before [16] using the implementation in the psych package Version 2.4.6 (https://CRAN.R-project.org/package=psych) [45]. Locally Estimated Scatterplot Smoothing (LOESS) was used to augment scatter plots with nonlinear, non-parametric regression curves [11], using the implementation in ggplot2 [51].

## Results

In total 97 consecutive patients were included into the analysis. 59 cases (60.8%) were contributed from one study center. The largest share of patients (72/96, 75.0%) accounted for definite MD (dMD). Mean age of all patients was 56.2 ± 15.0 years. Age and affected side were distributed evenly between diagnostic categories. Within the total cohort, the majority of cases were female (56.7%), with a borderline significant difference between diagnostic categories. The majority of patients (50/97, 51.6%) were users of hearing aids. Hearing aid use was significantly higher in the diagnostic category dMD (**Table 1**).

**Table 1:** Patient’s characteristics by diagnostic categories

| Characteristic | dMD, N = 72 (74%)<sup>1</sup> | pMD, N = 14 (14%)<sup>1</sup> | MC, N = 11 (11%)<sup>1</sup> | p-value<sup>2</sup> |
| --- | --- | --- | --- | --- |
| Study center |  |  |  | 0.3 |
| Greifswald | 29 (40%) | 7 (50%) | 2 (18%) |  |
| Munich | 43 (60%) | 7 (50%) | 9 (82%) |  |
| Affected side |  |  |  | 0.5 |
| left | 42 (58%) | 6 (43%) | 5 (45%) |  |
| right | 30 (42%) | 8 (57%) | 6 (55%) |  |
| Sex |  |  |  | **0.050** |
| m | 34 (47%) | 2 (14%) | 6 (55%) |  |
| f | 38 (53%) | 12 (86%) | 5 (45%) |  |
| Age [yrs] | 57 (50, 66) | 57 (54, 70) | 56 (43, 60) | 0.6 |
| Hearing aid user | 43 (61%) | 3 (21%) | 4 (36%) | **0.012** |
| missing | 2 | 0 | 0 |  |

<sup>1</sup>n (%);
<sup>2</sup>Fisher's exact test; Kruskal-Wallis rank sum test; Pearson's Chi-squared test;
Canonical diagnostic categories dMD, definite Menière's disease; pMD, probable Menière's disease [31]; experimental category MC, Menière's Characteristics [25].

Hearing loss as assessed by pure tone audiometry was asymmetric and worse in the affected ears with a mean 4PTA of 51.9 ± 30.2 dB HL (**Figure 1A**) compared to 19.3 ± 15.0 dB HL on the non-affected ears (**Figure 2B**). Mean difference between affected and unaffected ears was 32.6 ± 25.2 dB. 70 cases (72.2%) had an interaural difference of more than 15 dB, 51 (52.6%) an interaural difference of even more than 30 dB. Speech audiometry revealed severely impaired speech recognition in affected ears with a recognition score for monosyllables of 33.9% ± 42.9%. In contrast, unaffected ears achieved 86.6% ± 21.6%. There was a strong sigmoid correlation between 4PTA and speech recognition score (*p*<0.01) in a logistic regression model (**Figure 1C**). A breakdown of audiometric data by diagnostic categories revealed that patients from dMD showed significantly worse hearing when assessed by pure tone as well as speech audiometry **(Table 2)**.

**Figure 1:** Pure tone audiometry and speech recognition results Pure tone thresholds of all individual patients on affected (A) and non-affected (B) ears. Regression analysis of a correlation between average pure tone thresholds and speech recognition (C).

**Figure 2:** Distribution of SSQ-results within subscales and additional questionsAbsolute count of values in the investigated cohort.

**Table 2:** Audiometry by diagnostic categories

| **Test and side** | **dMD, N = 72<sup>1</sup>** | **pMD, N = 14<sup>1</sup>** | **MC, N = 11<sup>1</sup>** | ***p*-value<sup>2</sup>** |
| --- | --- | --- | --- | --- |
| 4PTA affected ear [dB HL] | 58 (39, 71) | 39 (10, 58) | 40 (19, 53) | **0.005** |
| 4PTA unaffected ear [dB HL] | 16 (9, 25) | 11 (8, 21) | 13 (8, 30) | 0.7 |
| speech recognition score affected ear [%] | 0 (0, 48) | 50 (10, 100) | 80 (0, 95) | **0.002** |
| speech recognition score unaffected ear [%] | 98 (90, 100) | 100 (95, 100) | 100 (85, 100) | 0.8 |

<sup>1</sup>Median (interquartile range);
<sup>2</sup>Fisher's exact test; Kruskal-Wallis rank sum test; Pearson's Chi-squared test;
Canonical diagnostic categories dMD, definite Menière's disease; pMD, probable Menière's disease [31]; experimental category MC, Menière's Characteristics [25].

The mean total score of the SSQ17 was 6.0 ± 2.1 (median 5.9, range 1.6-10.0). The mean score of the subscales was 5.3 ± 2.5 (median 5.4, range 0.5-10.0) for speech hearing, 5.1 ± 2.7 (median 4.6, range 0.0-10.0) for spatial hearing, and 7.2 ± 2.1 (median 7.8, range 1.2-10.0) for qualities of hearing.

Additional questions 1 and 2 yielded a mean of 8.2 ± 2.0 (median 9.0, range 1.8-10.0) and 6.1 ± 2.9 (median 7.0, range 0.0-10.0), respectively. Analysis of SSQ17-results by diagnostic category showed no statistically significant differences between categories, neither for the total score nor for domains or additional questions (**Table 3**). We further analyzed the distribution of responses for the subscales as well as the additional questions of the SSQ17. For the subscales as well as additional question 2, we did not notice floor or ceiling effects. A ceiling effect in additional question 1 signified that most individuals indicated good performance in the comparably simple listening situation with one conversational partner in a quiet room without reverberation (**Figure 2**).

**Table 3:** Results of the SSQ17 by diagnostic categories

|  | **dMD, N = 72<sup>1</sup>** | **pMD, N = 14<sup>1</sup>** | **MC, N = 11<sup>1</sup>** | **p-value<sup>2</sup>** |
| --- | --- | --- | --- | --- |
| total score | 5.9 (4.2, 7.5) | 7.2 (4.1, 9.4) | 5.8 (4.5, 7.8) | 0.7 |
| missing | 8 | 3 | 2 |  |
| speech hearing subscale | 5.0 (3.0, 7.0) | 6.2 (3.9, 8.4) | 6.3 (3.6, 7.8) | 0.3 |
| missing | 6 | 2 | 1 |  |
| spatial hearing subscale | 4.5 (2.8, 6.9) | 4.3 (2.8, 7.7) | 5.0 (4.2, 8.8) | 0.5 |
| missing | 3 | 0 | 0 |  |
| qualities of hearing subscale | 8.0 (6.3, 8.9) | 7.0 (4.6, 9.1) | 7.2 (4.8, 9.0) | 0.8 |
| missing | 0 | 0 | 0 |  |
| additional question 1: quiet room | 9.0 (8.0, 10.0) | 9.0 (8.0, 9.5) | 8.0 (7.0, 10.0) | 0.9 |
| missing | 1 | 1 | 1 |  |
| additional question 2: conversational effort | 6.0 (3.0, 8.1) | 8.0 (7.0, 9.5) | 6.5 (3.0, 8.0) | 0.057 |
| missing | 1 | 1 | 1 |  |

<sup>1</sup>n (%), Median (IQR);
<sup>2</sup>Fisher's exact test; Kruskal-Wallis rank sum test; Pearson's Chi-squared test;
Canonical diagnostic categories dMD, definite Menière's disease; pMD, probable Menière's disease [31]; experimental category MC, Menière's Characteristics [25].

To assess the applicability of the SSQ17 for this particular patient cohort with MD, we calculated Cronbach’s alpha for the total score and for the subscales. For the total score, Cronbach’s alpha was 0.960 (95% CI 0.95; 0.97). Speech hearing showed 0.936 (95% CI 0.91; 0.95), spatial hearing 0.956 (95% CI 0.94; 0.97), and qualities of hearing 0.904 (95% CI 0.87; 0.93). Thereby, Cronbach’s alpha resulted in excellent values for the total score and all three subscales, affirming the value of each individual item and validating the applicability to a patient cohort with Menière’s Disease. A breakdown of the consistency analysis to subscales and single items is given in **Table 4**. We also assessed correlations between subscales and additional questions. Statistically significant strong positive correlations were found between speech hearing and spatial hearing with *r*=0.755, qualities of hearing with *r*=0.706 as well as additional question 2 with *r*=0.705. All other pairwise correlations were positive as well but less pronounced.

**Table 4:**
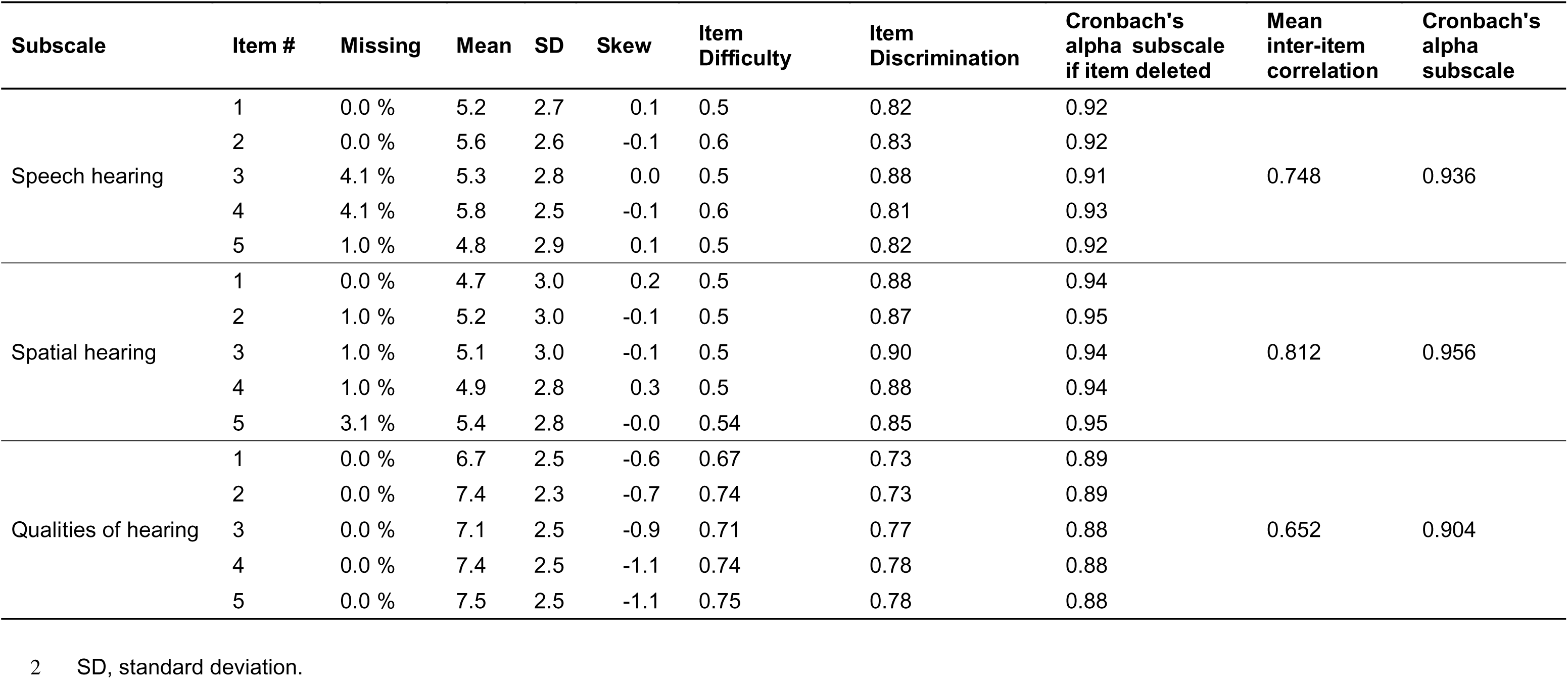
Internal consistency of the multi-item subscales of the SSQ17

Finally, we explored a correlation between audiometric variables as well as SSQ17 subscales and the additional questions. As higher thresholds in pure tone audiometry are a measure of more severe hearing impairment, we calculated correlations between SSQ17 and 4PTA on the affected ear. Excluding two outliers with very large 4PTA, there were statistically significant moderate to low negative correlations of responses in all three subscales as well as additional question 2 with pure tone thresholds of the affected ear (**Figure 3**). Correlations of speech recognition scores and SSQ are visually less clear due to an accumulation of observations at 0% speech recognition. However, there were statistically significant moderate positive relationships for the scales speech hearing, spatial hearing as well as the additional question 2 with speech recognition score on the affected ear (**Figure 4**).

**Figure 3:** Correlation between SSQ and average pure-tone thresholds A-C subscales; D and E additional questions; blue line: LOESS smoother, R - Spearman correlation coefficient.

**Figure 4:** Correlation between SSQ and speech recognition scores A-C subscales; D and E additional questions; blue line: LOESS smoother, R - Spearman correlation coefficient.

## Discussion

This is the first study to analyze results with a version of the SSQ in a large cohort of MD-patients. We found the SSQ17 being a viable tool to assess hearing disability in MD-patients. This was documented by the SSQ i) showing excellent internal consistency calculated by Cronbach’s alpha; ii) correlating negatively with pure tone thresholds (all subscales and additional question 2) and finally iii) correlating positively with speech recognition scores (subscales speech hearing and spatial hearing as well as additional question 2) in this cohort.

Pertaining to hearing assessment in MD patients, most studies utilized diagnostic tests like pure tone and speech audiometry. Analysis of SSQ results in MD patients, however, has been only done in the follow-up after cochlear implantation so far [40,47,54], and is therefore this study’s unique earmark. As we found an association between results in audiometry and the SSQ, this underlines the value of assessing both clinical and subjective hearing impairment for a comprehensive assessment of the sequelae of MD on hearing. The demographic characteristics of our population of 97 individuals with a mean age of 56.2 ± 15.0 years and 57% females are within the range of previous reports on large cohorts of patients with MD [5,35,53].

The SSQ is an established tool, besides the Abbreviated Profile of Hearing Aid Benefit (APHAB), as one of the two primary measures to assess the effect of hearing devices on hearing-related quality of life [29]. Moreover, the SSQ shows particular sensitivity when it comes to evaluating the benefit with hearing devices in advanced listening conditions like speech perception in noise and sound localization [28]. Age-matched comparison seems to be of relevance, since when looking at other studies’ scores, it is suggested that higher SSQ-scores are found in younger individuals and *vice versa* [1]. When comparing the SSQ values from our MD cohort to an age-matched sample of presumably normal hearing adults (age 50-59 years) who exhibited a total score of 8.1 (subscales speech: 7.6; spatial 8.0; qualities: 9.0), our MD patients revealed worse scores below the 0.1 quantile of SSQ results of this reference population at the given age [50]. Regarding a general definition of a threshold for disability, the World Health Organization’s International Classification of Functioning, Disability and Health suggests a general cut-of value of 2 standard deviations below the mean of an unimpaired population [13]. A consecutive study determined this threshold for the SSQ in 11 adult normal hearing-individuals, resulting in values of 7.25 for the total score as well as 6.84 for the speech hearing, 6.14 for the spatial hearing, and 8.18 for the qualities of hearing subscale [49]. Applying this definition, this definition, the values observed in our population of MD-patients of 6.0 for the total score as well as 5.3, 5.1, and 7.2 for the respective subscales, are well within the range of a pathologic hearing condition.

For hearing impaired individuals without hearing aids, asymmetric hearing was reported to result in severe impairment of hearing-related quality of life as assessed by the SSQ: already Nobel and Gatehouse utilized the SSQ questionnaire to consider the status of hearing loss on both ears. Comparing our results to a group of individuals with bilaterally symmetric hearing impairment, the SSQ total score was slightly higher (our MD patients: 6.0 ±2.1 vs. 5.9). Moreover, it was profoundly higher when compared to individuals with asymmetric hearing loss (4.7) [37]. However, for that cohort the underlying cause for asymmetric hearing was not documented, therefore a sound comparison to our homogenous population of MD patients is limited. It is important to consider the fluctuating nature of aural symptoms in MD. Thereby, any given assessment might only represent a snapshot of the state of hearing, whereas patients with a different cause for their asymmetric hearing will most likely have a stable or more maintained level of hearing loss for a long time. In 49 patients with asymmetric hearing loss of mixed etiology, significant differences were found in the total score and all subscales of the SSQ when compared to the values of 11 normal-hearing individuals from a control group [49]. Thereby, 75% of those AHL-patients exhibited a hearing disability in this study when the definition of Demeester and coworkers of 2 standard deviations below the mean value of normal listeners [13] is applied. While this underlines the usefulness of the SSQ in asymmetric hearing loss, neither this nor any other patient-reported outcome instrument has been designed specifically for asymmetric hearing loss so far.

Other MD studies assessed performance with cochlear implant (CI) with the SSQ. Comparing the values of our obviously better hearing cohort with their values, our patients would be found at the upper range of those previous reports (other studies: 4.0 ±1.5 at 12 months [47]; 6.1 at 12 months [54]). The MD cohort of Wrobel et al. included both individuals with uni- and bilateral hearing loss, as well as uni- and bilateral CI users and therefore does not allow to estimate the effect of the typically asymmetric characteristic of hearing loss in mostly unilaterally affected MD-patients [54]. The study of Perkins et al. reported values of 3 MD-patients with labyrinthectomy and concurrent CI [40]: performance with CI in this specific MD group is most likely determined by the initial incentive for this specific treatment concept. Patients receiving a labyrinthectomy proceed to this deliberate step of treatment because of their vertiginous symptoms. Hence, hearing with the CI may be secondary and motivation to train for their performance with the CI might be lacking. In addition to that, this particular patient group qualifies as single sided deafness, another known complex group of CI patients to motivate for their hearing training due to their contralateral normal hearing ear.

Beyond MD, a wide range of SSQ-scores has been reported from populations with different etiologies of hearing loss. In most cases, the SSQ was employed to assess the benefit of an intervention, like the provision of hearing aids [10,23]. Adapting their values for comparison to the presented study, results for the subscales were in keeping with unaided hearing-impaired individuals. That being said, however, our cohort represents a mix regarding hearing rehabilitation with a slight majority (50/97, 51.6%) of hearing aid users. Thereby, variability in the general properties of reported cohorts has to be considered. This might, on the other hand, influence the latent variables that may be assessed by the SSQ. The initial English version of the SSQ contains 49 items and interestingly, a factor analysis in 1220 participants with mostly symmetrical hearing loss (in three groups representing unaided, unilaterally aided and bilaterally aided) showed three factors that can approximately be related to the three subscales. A fourth factor, associated with effort, is only implied in this dataset [3]. While we suppose that effort plays a considerable role in the hearing experience of MD-patients, we did not perform a factor analysis in this study. This is an interesting question for a follow-up-analysis, in particular as MD-patients with asymmetric hearing loss have not been studied so far in this regard. However, we expect this analysis to require a higher number of cases. Therefore, it is scheduled as a future analysis within the multicenter prospective patient registry SEMM.

Internal consistency of the SSQ has not yet been assessed in a population of MD patients. With Cronbach’s alpha of 0.960 for the total score and 0.904 to 0.956 in the subscales, we see the SSQ as a reliable instrument both in total and with regard to the subscales and their implied latent variables. For the English version, a threshold of 0.9 has been suggested for a clinical applicability for the instrument in whole as well as for the three subscales individually [9,48]. For the Dutch version with 50 items, Cronbach’s alpha was determined at 0.96 for a subset of 45 items as well as 0.93, 0.95, and 0.92 for the speech, spatial, and quality domains, respectively [13]. Cronbach’s alpha was reported to be 0.83 for the 5-item Dutch SSQ5 [13], 0.93 for an English 12-item SSQ12 [38], and 0.94 for the German SSQ17 also used in the present study [27] as well as for a French 15-item SSQ [36]. Looking at these studies, the German language 17-item SSQ shows comparably good internal consistency for MD-patients as it does for different etiologies of hearing loss.

While being applicable to MD, the SSQ is limited to assessing hearing-related disability. From the patient perspective, however, debilitating vertigo spells are a different and major facet of disease burden. This has been described by an assessment of illness intrusiveness in 74 cases with MD, where vertigo had the greatest detrimental effect on daily life, followed by hearing loss and tinnitus [5]. The dizziness-specific quality of life instrument Dizziness Handicap Inventory illustrates a mild handicap in MD-patients, again with a statistically significant worse value for bilateral cases [5,32]. With the Vertigo Symptom Scale, Yardley and coworkers described vertigo severity as the most prominent factor for impaired generic quality of life in a community-based sample of individuals with MD [55].

Furthermore, overall health-related quality of life is also severely impaired in MD. Compared to other disorders, MD symptoms seem to carry high intrusiveness in general [5]. It is important to note that symptom-specific instruments will always only capture a specific aspect of overall quality of life. Generic instruments for health-related quality of life employed for MD in the past included the Short Form 36 (SF36) Health Instrument. This also showed a considerable burden in MD with impairment of the quality of life on a similar level as other severe chronic conditions [55]. An additional effect to already reduced quality of life has been described in bilateral cases of MD [32]. Patient-reported outcome measures established specifically for MD may be a practical solution but risk losing detail compared to the assessment of advanced hearing conditions by the SSQ. Disease-specific tools include the Menière’s Disease Outcome Questionnaire [7] and as a recent addition the Menière’s Disease Quality of Life (MenQOL) [42]. Both instruments are not yet available in a validated German-language version. The “Ménière’s Disease Patient-Oriented Symptom-Severity Index” [5,19] has recently been validated in German [41] and might in the future be a promising option in this regard.

## Conclusion

Patient-reported outcome measures hold value in managing conditions which involve perception-based symptoms. This study found the SSQ being a viable tool to assess burden of hearing loss in advanced listening conditions in patients with Meniere’s disease, thereby being a potential addition to current clinical diagnostic tools.

## Acknowledgements

The authors are grateful to the clinical and technical team involved at the dizziness clinics in Munich and Greifswald. <u>The doctoral thesis of Dr. Irina Adler</u> is based on parts of the data of this work. Preliminary results of this work were presented on May 11^th^, 2024 at the 95^th^ Annual Meeting of the German Society of Oto-Rhino-Laryngology, Head and Neck Surgery in Essen (https://doi.org/10.1055/s-0044-1784278) and on May 30^th^, 2025 at the 96^th^ Annual Meeting of the German Society of Oto-Rhino-Laryngology, Head and Neck Surgery in Frankfurt am Main.

## Data availability

The data that supports the findings of this study cannot be made openly available due to national data protection laws. However, upon individual reasonable request the author team will provide an anonymized dataset for scientific purposes only.

## References

1. M. Adel Ghahraman, M. Ashrafi, G. Mohammadkhani, and S. Jalaie, Effects of aging on spatial hearing, Aging Clin Exp Res 32 (2020), 733–739.

2. Y. Agrawal, K.G. Pineault, and Y.R. Semenov, Health-related quality of life and economic burden of vestibular loss in older adults, Laryngoscope Investig Otolaryngol 3 (2018), 8–15.

3. M.A. Akeroyd, F.H. Guy, D.L. Harrison, and S.L. Suller, A factor analysis of the SSQ (Speech, Spatial, and Qualities of Hearing Scale), Int J Audiol 53 (2014), 101–14.

4. T.H. Alexander and J.P. Harris, Current epidemiology of Meniere’s syndrome, Otolaryngol Clin North Am 43 (2010), 965–70.

5. M. Arroll, C.P. Dancey, and E.A. Attree, People With Symptoms of Menière’s Disease: The Relationship Between Illness Intrusiveness, Illness Uncertainty, Dizziness Handicap, and Depression, 33 (2012).

6. C.D. Balaban, R.G. Jacob, and J.M. Furman, Neurologic bases for comorbidity of balance disorders, anxiety disorders and migraine: neurotherapeutic implications, Expert Rev Neurother 11 (2011), 379–94.

7. D.P. Ballard, D.C. Sukato, A. Timashpolsky, S.C. Babu, R.M. Rosenfeld, and M. Hanson, QualityloflLife Outcomes following Surgical Treatment of Ménière’s Disease: A Systematic Review and Metalanalysis, Otolaryngol Neck Surg 160 (2019), 232–238.

8. G.J. Basura, M.E. Adams, A. Monfared, S.R. Schwartz, P.J. Antonelli, R. Burkard, M.L. Bush, J. Bykowski, M. Colandrea, J. Derebery, E.A. Kelly, K.A. Kerber, C.F. Koopman, A.A. Kuch, E. Marcolini, B.J. McKinnon, M.J. Ruckenstein, C.V. Valenzuela, A. Vosooney, S.A. Walsh, L.C. Nnacheta, N. Dhepyasuwan, and E.M. Buchanan, Clinical Practice Guideline: Ménière’s Disease, Otolaryngol Neck Surg 162 (2020).

9. J.M. Bland and D.G. Altman, Statistics notes: Cronbach’s alpha, BMJ 314 (1997), 572–572.

10. M. Boymans and W.A. Dreschler, Audiologist-Driven Versus Patient-Driven Fine Tuning of Hearing Instruments, Trends Amplif 16 (2012), 49–58.

11. W.S. Cleveland, Robust Locally Weighted Regression and Smoothing Scatterplots, J Am Stat Assoc 74 (1979), 829–836.

12. L.J. Cronbach, Coefficient alpha and the internal structure of tests, Psychometrika 16 (1951), 297– 334.

13. K. Demeester, V. Topsakal, J.-J. Hendrickx, E. Fransen, L. Van Laer, G. Van Camp, P. Van De Heyning, and A. Van Wieringen, Hearing Disability Measured by the Speech, Spatial, and Qualities of Hearing Scale in Clinically Normal-Hearing and Hearing-Impaired Middle-Aged Persons, and Disability Screening by Means of a Reduced SSQ (the SSQ5), Ear Hear 33 (2012), 615–616.

14. N.Y. Dwyer, J.B. Firszt, and R.M. Reeder, Effects of unilateral input and mode of hearing in the better ear: self-reported performance using the speech, spatial and qualities of hearing scale, Ear Hear 35 (2014), 126–36.

15. B.F. van Esch, H. van der Zaag-Loonen, T. Bruintjes, T. Kuijpers, and P.P.G. van Benthem, Interventions for Menière’s disease: an umbrella systematic review, BMJ Evid-Based Med 27 (2022), 235–245.

16. L.S. Feldt, D.J. Woodruff, and F.A. Salih, Statistical Inference for Coefficient Alpha, Appl Psychol Meas 11 (1987), 93–103.

17. P.C. Fernandes, B. Takegawa, F.F. Ganança, and D. Gil, Speech Perception in Ménière Disease, Int Arch Otorhinolaryngol 27 (2023), 613–619.

18. S. Gatehouse and W. Noble, The Speech, Spatial and Qualities of Hearing Scale (SSQ), Int J Audiol 43 (2004), 85–99.

19. G.A. Gates and A.M. Verrall, Validation of the Ménière’s Disease Patient-Oriented Symptom-Severity Index, Arch Otolaryngol Neck Surg 131 (2005), 863.

20. C. Gini, Variabilita e Mutabilita contributo allo studio delle distribuzioni e delle relazioni statistiche, Tipografia di Paolo Cuppini, Bologna, 1912.

21. K.H. Hahlbrock, [Speech audiometry and new word-tests], Arch Ohren Nasen Kehlkopfheilkd 162 (1953), 394–431.

22. D.A. Hanes and G. McCollum, Cognitive-vestibular interactions: a review of patient difficulties and possible mechanisms, J Vestib Res Equilib Orientat 16 (2006), 75–91.

23. S.S. Houmøller, A. Wolff, L.-T. Tsai, S.K. Narayanan, D.D. Hougaard, M.L. Gaihede, T. Neher, C. Godballe, and J.H. Schmidt, Impact of hearing aid technology level at first-fit on self-reported outcomes in patients with presbycusis: a randomized controlled trial, Front Aging 4 (2023), 1158272.

24. Y. Huang, P. Zhao, Z. Han, J. Xie, Y. Liu, S. Gong, and Z. Wang, Evaluation of the relationship between endolymphatic hydrops and hearing loss in Meniere’s disease based on three-dimensional real inversion recovery sequence, Braz J Otorhinolaryngol 89 (2023), 101314.

25. F. Ihler, I. Stoycheva, J.L. Spiegel, D. Polterauer, J. Müller, R. Strobl, and E. Grill, Diagnosis of Menière’s disease according to the criteria of 2015: Characteristics and challenges in 96 patients, J Vestib Res Equilib Orientat 32 (2022), 271–283.

26. S. Iwasaki, H. Sano, S. Nishio, Y. Takumi, M. Okamoto, S. Usami, and K. Ogawa, Hearing handicap in adults with unilateral deafness and bilateral hearing loss, Otol Neurotol Off Publ Am Otol Soc Am Neurotol Soc Eur Acad Otol Neurotol 34 (2013), 644–9.

27. J. Kießling, L. Grugel, H. Meister, and M. Meis, German translations of questionnaires SADL, ECHO and SSQ and their evalution, GMS Z Für Audiol – Audiol Acoust 50 (2011), 6–16.

28. P.T. Kitterick, L. Lucas, and S.N. Smith, Improving Health-Related Quality of Life in Single-Sided Deafness: A Systematic Review and Meta-Analysis, Audiol Neurotol 20 (2015), 79–86.

29. P.T. Kitterick, S.N. Smith, and L. Lucas, Hearing Instruments for Unilateral Severe-to-Profound Sensorineural Hearing Loss in Adults: A Systematic Review and Meta-Analysis, Ear Hear 37 (2016), 495–507.

30. D.P. Kumpik and A.J. King, A review of the effects of unilateral hearing loss on spatial hearing, Hear Res 372 (2019), 17–28.

31. J.A. Lopez-Escamez, J. Carey, W.-H. Chung, J.A. Goebel, M. Magnusson, M. Mandalà, D.E. Newman-Toker, M. Strupp, M. Suzuki, F. Trabalzini, and A. Bisdorff, Diagnostic criteria for Menière’s disease, J Vestib Res Equilib Orientat 25 (2015), 1–7.

32. J.A. Lopez-Escamez, D. Viciana, and P. Garrido-Fernandez, Impact of Bilaterality and Headache on Health-Related Quality of Life in Meniere’s Disease, (n.d.).

33. M.F.J.A. van der Lubbe, A. Vaidyanathan, V. van Rompaey, A.A. Postma, T.D. Bruintjes, D.M. Kimenai, P. Lambin, M. van Hoof, and R. van de Berg, The “hype” of hydrops in classifying vestibular disorders: a narrative review, J Neurol 267 (2020), 197–211.

34. L. Lucas, R. Katiri, and P.T. Kitterick, The psychological and social consequences of single-sided deafness in adulthood, Int J Audiol 57 (2018), 21–30.

35. M.D.C. Moleon, L. Torres-Garcia, A. Batuecas-Caletrio, N. Castillo-Ledesma, R. Gonzalez-Aguado, L. Magnoni, M. Rossi, F. Di Berardino, V. Perez-Guillen, G. Trinidad-Ruiz, and J.A. Lopez-Escamez, A Predictive Model of Bilateral Sensorineural Hearing Loss in Meniere Disease Using Clinical Data, Ear Hear 43 (2022), 1079–1085.

36. A. Moulin, J. Vergne, S. Gallego, and C. Micheyl, A New Speech, Spatial, and Qualities of Hearing Scale Short-Form: Factor, Cluster, and Comparative Analyses, Ear Hear 40 (2019), 938– 950.

37. W. Noble and S. Gatehouse, Interaural asymmetry of hearing loss, Speech, Spatial and Qualities of Hearing Scale (SSQ) disabilities, and handicap, Int J Audiol 43 (2004), 100–14.

38. W. Noble, N.S. Jensen, G. Naylor, N. Bhullar, and M.A. Akeroyd, A short form of the Speech, Spatial and Qualities of Hearing scale suitable for clinical use: the SSQ12, Int J Audiol 52 (2013), 409–12.

39. F. Orji, The Influence of Psychological Factors in Meniere’s Disease, Ann Med Health Sci Res 4 (2014), 3–7.

40. E. Perkins, M. Rooth, M. Dillon, and K. Brown, Simultaneous labyrinthectomy and cochlear implantation in unilateral meniere’s disease, Laryngoscope Investig Otolaryngol 3 (2018), 225– 230.

41. M. Plath, M. Sand, M. Appel, S. Euteneuer, M. Praetorius, I. Baumann, and K. Zaoui, Validity of the German Menière’s Disease Patient-Oriented Symptom Severity Index, Laryngo-Rhino-Otol 102 (2023), 856–866.

42. A.E. Quimby, J.A. Brant, J.P. Staab, and M.J. Ruckenstein, Development and Initial Validation of a Meniere’s Disease Quality of Life Instrument: The MenQOL, The Laryngoscope 134 (2024), 4351–4357.

43. R Core Team, R: A Language and Environment for Statistical Computing, (2024).

44. E. Ravin, A.E. Quimby, M. Bartellas, S. Swanson, T.P. Hwa, D.C. Bigelow, J.A. Brant, and M.J. Ruckenstein, An Update on the Epidemiology and Clinicodemographic Features of Meniere’s Disease, The Laryngoscope 134 (2024), 3310–3315.

45. W. Revelle, psych: Procedures for Psychological, Psychometric, and Personality Research, (2024).

46. H.G. Rizk, N.K. Mehta, U. Qureshi, E. Yuen, K. Zhang, Y. Nkrumah, P.R. Lambert, Y.F. Liu, T.R. McRackan, S.A. Nguyen, and T.A. Meyer, Pathogenesis and Etiology of Ménière Disease: A Scoping Review of a Century of Evidence, JAMA Otolaryngol Neck Surg 148 (2022), 360.

47. I. Sanchez-Cuadrado, M. Calvino, J.M. Morales-Puebla, J. Gavilán, T. Mato, J. Peñarrocha, M.P. Prim, and L. Lassaletta, Quality of Life Following Cochlear Implantation in Patients With Menière’s Disease, Front Neurol 12 (2021), 670137.

48. G. Singh and M. Kathleen Pichora-Fuller, Older adults’ performance on the speech, spatial, and qualities of hearing scale (SSQ): Test-retest reliability and a comparison of interview and self-administration methods, Int J Audiol 49 (2010), 733–740.

49. N. Vannson, C. James, B. Fraysse, K. Strelnikov, P. Barone, O. Deguine, and M. Marx, Quality of Life and Auditory Performance in Adults with Asymmetric Hearing Loss, Audiol Neurotol 20 (2015), 38–43.

50. P. Von Gablenz, F. Otto-Sobotka, and I. Holube, Adjusting Expectations: Hearing Abilities in a Population-Based Sample Using an SSQ Short Form, Trends Hear 22 (2018), 2331216518784837.

51. H. Wickham, ggplot2, Springer International Publishing, Cham, 2016.

52. O.B. Wie, A.H. Pripp, and O. Tvete, Unilateral deafness in adults: effects on communication and social interaction, Ann Otol Rhinol Laryngol 119 (2010), 772–81.

53. P. WladislavoskylWaserman, G.W. Facer, B. Mokri, and L.T. Kurland, Meniere’s disease: A 30lYear epidemiologic and clinical study in Rochester, MN, 1951l1980., The Laryngoscope 94 (1984), 1098–1102.

54. C. Wrobel, N.F. Bevis, A. Klinge-Strahl, N. Strenzke, and D. Beutner, Performance and self-perceived hearing impairment after cochlear implantation in Menière’s disease, Laryngoscope Investig Otolaryngol 7 (2022), 219–225.

55. L. Yardley, B. Dibb, and G. Osborne, Factors associated with quality of life in Menière’s disease, Clin Otolaryngol Allied Sci 28 (2003), 436–41.

56. S. Yitzhaki, The GMD: A superior Measure of Variability for Non-Normal Distributions, SSRN Electron J (2002).

